# Strengthening the translation of malaria modelling into policy: Design, implementation, and early outcomes of the Regional Malaria Modelling Translational Fellowship

**DOI:** 10.64898/2026.08.04.26359706

**Authors:** Sheetal P Silal, Rachel A Hounsell, Sadiq Wanjala

## Abstract

Malaria programmes increasingly rely on modelled evidence to support intervention prioritisation, resource allocation, and elimination planning, yet a persistent gap remains between technical modelling outputs and their use in decision-making. We describe the design, implementation, and early outcomes of the Regional Malaria Modelling Translational Fellowship, a six-month executive education programme delivered in 2025 to 21 fellows nominated by national malaria programmes in seven African countries. The Fellowship was designed to strengthen translational capacity by focusing on question formulation, systems thinking, model design and critique, interpretation of outputs, uncertainty, health economics, communication, and stakeholder engagement. The hybrid structure combined three intensive in-person blocks with regular virtual sessions. Country-teams work on capstone projects throughout the Fellowship, applying learnings to develop policy-relevant modelling proposals aligned with national malaria priorities. The programme was accredited as a University of Cape Town short course, which supported credibility, participant commitment, and institutional endorsement. Early evaluation showed improvements across all competency domains, with the largest gains in fellows’ confidence in applying modelling to decision-making, translating model findings into recommendations, and communicating technical results to non-technical audiences. Qualitative feedback suggested that the Fellowship helped shift participants’ engagement with modelling from passive acceptance of results toward critical interpretation, collaborative dialogue, and practical application. These findings suggest that translational, executive-style training can strengthen the interface between modelling and malaria policy. The publicly available curriculum offers a replicable framework that may be adapted for other infectious disease and public health settings where modelling evidence is increasingly central to decision-making.

**Highlights:**

- Strengthened national malaria programmes’ capacity to use modelled evidence.
- An accredited six-month fellowship trained 21 decision-makers from seven countries.
- Country-led capstone projects applied learning to national malaria priorities.
- Fellows gained confidence in interpreting and communicating model outputs.
- The open curriculum offers a replicable model for other disease programmes.

## 1. Introduction

Malaria remains a persistent and evolving public health challenge, placing a significant burden on communities, health systems, and economies across endemic regions. While important progress has been made over the past two decades, gains have plateaued in many settings, and emerging threats highlight the urgent need for more effective, data-driven approaches to accelerate control and elimination [1]. To effectively address the malaria burden and achieve the goal of elimination, it is imperative to have robust, evidence-based strategies and policies [2]. Disease modelling plays a crucial role in informing decision-making by providing insights into disease transmission and dynamics, evaluating the impact of different interventions, and identifying optimal strategies for malaria control and elimination [3,4]. However, the true value of modelling lies not only in its technical rigour, but in its translation into actionable policies and programmes that reflect local realities.

Realising this potential requires strong collaboration across a diverse set of stakeholders, including national malaria programmes (NMP), researchers and modellers, policymakers, funders, and implementing partners. Modelling is increasingly available, but decision-makers and users of modelled output may lack the confidence, time, vocabulary, skills, or institutional mechanisms to interpret and use outputs [5]. Beyond interpretation and use, it is essential that users of modelled output can assess the model’s validity and suitability to support the issue at hand and engage with the uncertainty produced by the models. Sustained engagement and dialogue are essential to ensure that modelling outputs are relevant, understood, and effectively used in decision-making.

In this paper, we describe the design, implementation, and early outcomes of the inaugural Regional Malaria Modelling Translational (RMMT) Fellowship, a six-month executive programme designed to strengthen the use of modelled evidence in malaria policy and programme decision-making across seven African countries. The RMMT Fellowship objectives are to 1) Bridge the gap between modelling and effective decision-making; 2) Strengthen capacity to interpret and utilise modelling outputs; 3) Build relationships with modellers and decision-makers for impactful collaboration; 4) Develop actionable recommendations tailored to countries’ needs; 5) Strengthen technical leadership to advance malaria control and elimination; and 6) Enhance advocacy for resource allocation using modelling and data-driven insights.

The inaugural cohort comprised 21 fellows nominated by their respective NMPs from Angola, Eswatini, Ghana, Mozambique, Namibia, South Africa, and Zambia. Fellows were nominated by their Ministries of Health and reflect a diverse cross-section of technical and leadership roles within malaria programmes, including programme directors, monitoring and evaluation specialists, surveillance officers, data managers, clinicians, and policy advisors. This diversity was a deliberate design feature of the Fellowship, recognising that effective translation of modelling into policy requires engagement across multiple functions within malaria programmes and the broader health system. The cohort brought together individuals with expertise spanning epidemiology, medicine, statistics, data science, information technology, and public health, many of whom are directly involved in strategic planning, programme implementation, surveillance and performance monitoring at national and sub-national levels.

Several fellows held senior leadership positions within Ministries of Health and NMPs, while others served critical technical roles in data management, surveillance, and operational research. All had extensive experience working with routine health data, implementing malaria interventions, and contributing to national strategies, positioning them well to apply modelling insights in decision-making processes. Fellows’ varied backgrounds and perspectives enriched peer learning and collaboration, while ensuring that modelling approaches explored during the Fellowship remained grounded in real-world programmatic needs.

## 2. Methods

### 2.1 Delivery structure

The inaugural RMMT Fellowship was delivered through a hybrid format to maximise contact time while fitting in with the demands of fellows’ work schedules (Figure 1). Three immersive, full-time in-person blocks, each a week, were held in May, August, and October 2025. In between blocks, virtual sessions of 90 minutes duration were held twice a month (nine in total). All costs for the delivery of the programme were sponsored by the funder.

**Figure 1.**
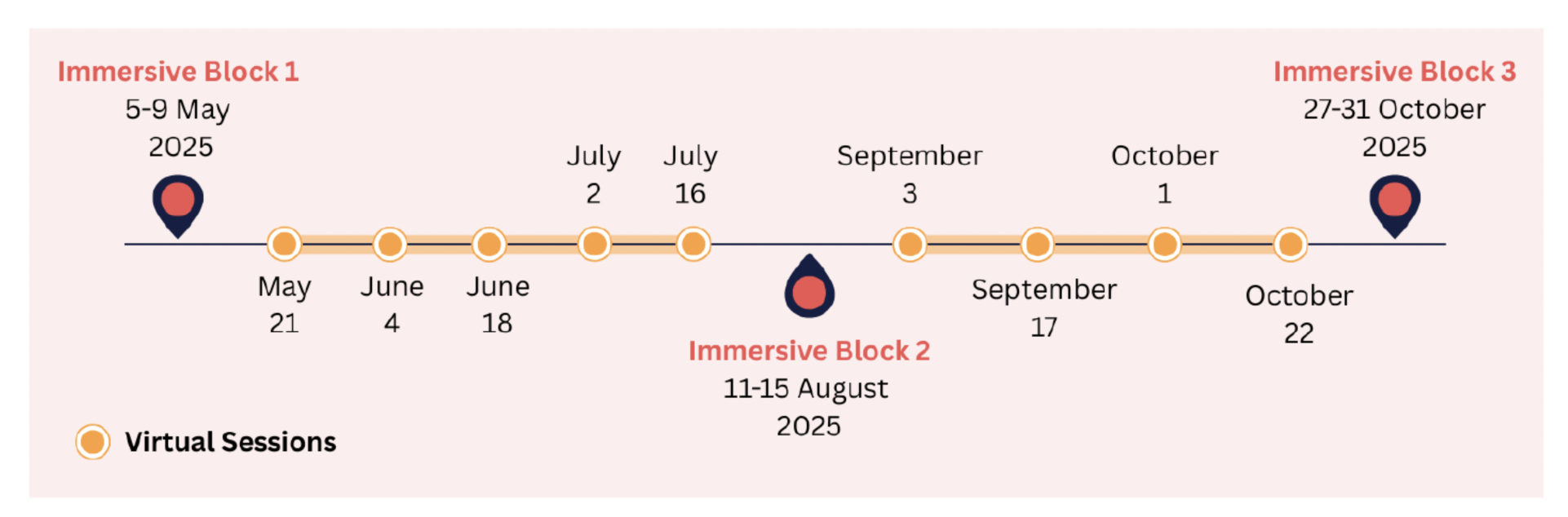
RMMT Fellowship course delivery structure

The RMMT Fellowship programme is an accredited University of Cape Town Short course (Translation of Modelling for Implementation) for which candidates received a certificate of completion on the basis of attendance for the three contact weeks and 80% of virtual sessions. All 21 fellows in the inaugural cohort successfully completed the programme.

### 2.2 Curriculum design

The curriculum was designed to provide a comprehensive and practical foundation in the use of modelling for malaria policy and decision-making. It spans the full modelling process—from framing policy-relevant questions and understanding systems, through model design, development, and critique, to interpreting outputs and communicating findings—while also addressing cross-cutting themes such as uncertainty, health economics, and stakeholder engagement. The selection of topics reflects the needs of the fellowship audience, balancing technical understanding with practical application to ensure participants can both engage critically with models and use their results in real-world contexts. The modules were structured to build on one another, reinforcing key concepts while allowing space for reflection and application. A mix of teaching approaches was used throughout, including lectures to introduce core concepts, facilitated discussions to draw on participant experience, and interactive and group-based exercises to apply learning to relevant scenarios, fostering both technical confidence and collaborative application skills.

### 2.3 Approach to learning

The RMMT Fellowship was designed as a practical, participant-centred learning programme founded on the following concepts:

- Its aim was not only to introduce modelling concepts, but to support fellows to apply modelling, evidence synthesis, and technical–policy dialogue to real malaria programme decisions.
- Modelling was presented as a collaborative process that depends on clear questions, contextual understanding, appropriate data, transparent assumptions, and effective communication.
- Learning activities were designed to help fellows move between concepts, application, and reflection. Modules combined short technical inputs with facilitated discussion, practical exercises, country team work, and opportunities to connect learning to capstone projects. This structure allowed fellows to test ideas, question assumptions, and consider how modelling can support decision-making in their own contexts.
- The learning approach recognises that uncertainty is central to both modelling and public health decision-making. Rather than treating uncertainty as a weakness, the curriculum used it as an opportunity to discuss confidence, risk, data limitations, and alternative scenarios. Fellows were supported to interpret uncertainty in ways that are meaningful for programme planning and policy decisions.

The Fellowship used an iterative learning model. Ideas introduced in early modules were revisited and strengthened through later sessions, practical exercises, and capstone project work. Fellows gradually built from understanding modelling concepts to formulating policy-relevant questions, interpreting outputs, communicating findings, and considering how modelling evidence can be embedded in programme decision-making.

### 2.4 Capstone projects

A central component of the RMMT Fellowship curriculum was the Capstone Project, through which fellows applied the knowledge and skills acquired during the programme to real-world policy challenges. Working collaboratively in country teams, fellows identified and developed proposals (Box 1) to answer a high-impact, policy-relevant question aligned with national and/or sub-national malaria priorities through a series of analyses that included modelling. Each Capstone Project was designed to empower the country team to develop and direct a modelling study rather than only consuming analyses in which they may have had minimal input. The process tested the fellows’ understanding of modelling and interpretation of model-based outputs and emphasised the practical application of modelling to inform intervention design, optimise resource allocation, and accelerate progress toward malaria control and elimination goals.

##### Box 1. Capstone Project Proposal Components

- Background and policy context, outlining the national malaria landscape and rationale for the selected priority question.
- Definition of the policy problem, clearly linking modelling needs to decision-making gaps.
- Research needs, incorporating both modelling and complementary non-modelling analyses required to inform the policy decision.
- Key modelling questions, spanning both health and economic perspectives.
- Data requirements, identifying necessary epidemiological, intervention, and cost data inputs and assessing the quality and availability of these inputs.
- Outcomes of interest, such as reductions in morbidity and mortality, progress toward elimination, and cost-effectiveness.
- Scenario analyses, exploring different intervention strategies, coverage levels, and implementation approaches.
- Economic evaluations, including cost and cost-effectiveness considerations to support resource allocation decisions.
- Illustrative model outputs and visualisations, used to communicate potential impacts and key policy messages.
- SWOT analysis, assessing the feasibility of implementing proposed strategies within each country’s context.
- Next steps, outlining pathways for further analysis, stakeholder engagement, and policy uptake.

During the final week of the Fellowship, country teams presented their Capstone Project proposals (Figure 2) to the full cohort and invited representatives from Ministries of Health and NMPs, who joined virtually. These presentations served as a platform for fellows to demonstrate the knowledge and skills gained throughout the programme, particularly their ability to translate modelling concepts into clear, policy-relevant recommendations. Importantly, engagement with key national stakeholders helped to validate the relevance of the proposed analyses, advocate for the practical value of modelling in informing programme decisions and demonstrate the gains of participation in the RMMT Fellowship. This interaction also laid the groundwork for continued collaboration and positioned the projects as a foundation for future modelling work within NMPs.

**Figure 2.**
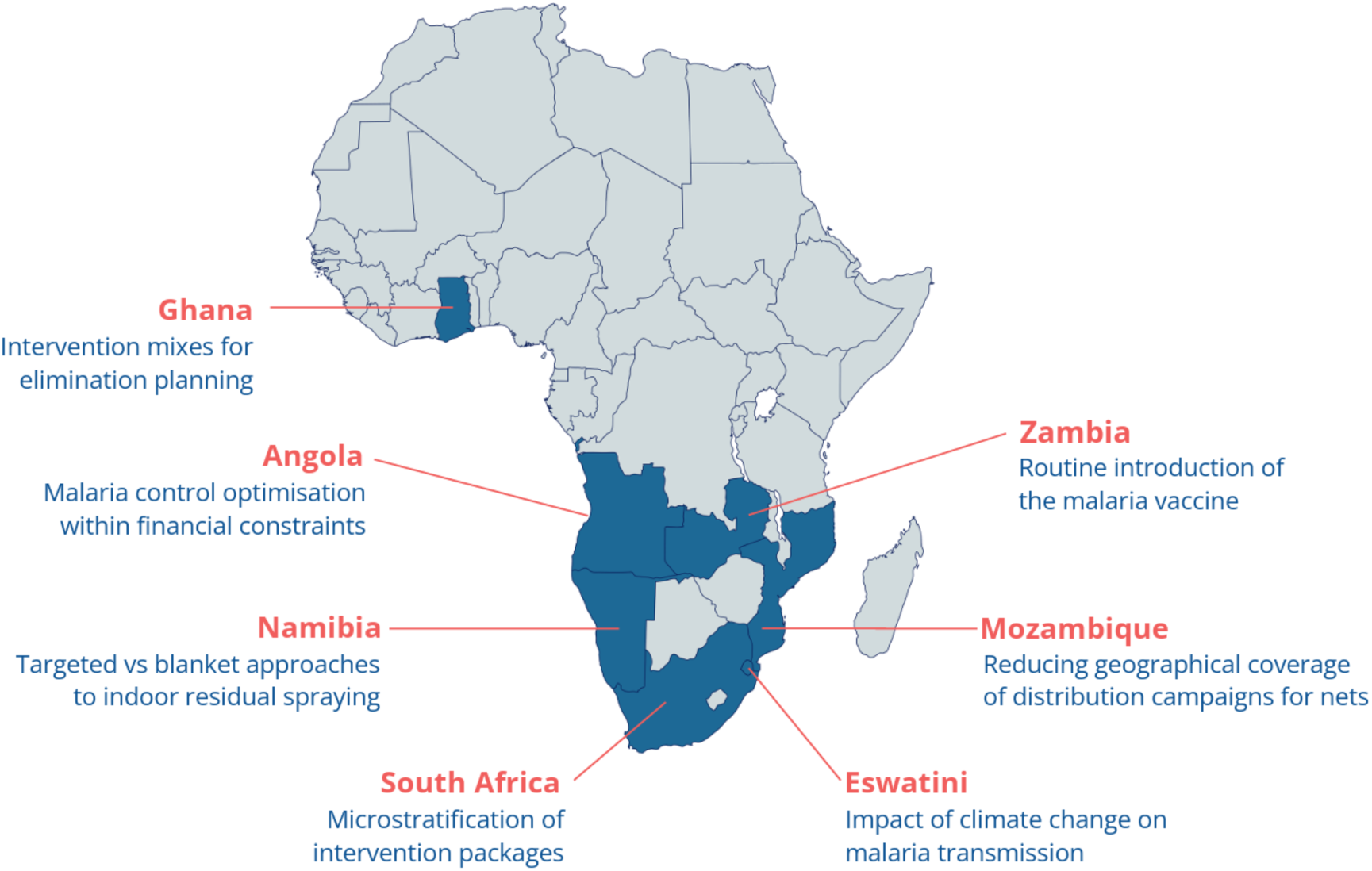
Capstone project topics on malaria control and elimination that were chosen by country teams during the 2025 RMMT Fellowship cohort

Collectively, the Capstone Projects demonstrated the value of embedding modelling within decision-making processes. They highlighted how locally led, policy-driven analyses can generate actionable insights to guide programme planning, improve allocative efficiency, and support evidence-based advocacy for sustained investment in malaria programmes.

### 2.5 Approach to monitoring and evaluation

We conducted a longitudinal assessment of the RMMT Fellowship, designed to track changes in fellows’ competencies and perceptions over the course of the programme. The monitoring and evaluation (M&E) was structured across three phases—baseline, midline, and final evaluations—each serving a distinct purpose, from establishing initial expectations and competencies to identifying areas for improvement and assessing overall learning outcomes. A mixed-methods approach was employed, combining quantitative analysis of a 14-item Likert scale across key thematic areas with qualitative insights gathered through interactive and open-ended feedback (Figure 3). Together, these methods provided a robust framework for understanding how fellows’ ability to engage with and translate modelling into policy-relevant action evolved throughout the Fellowship.

**Figure 3.**
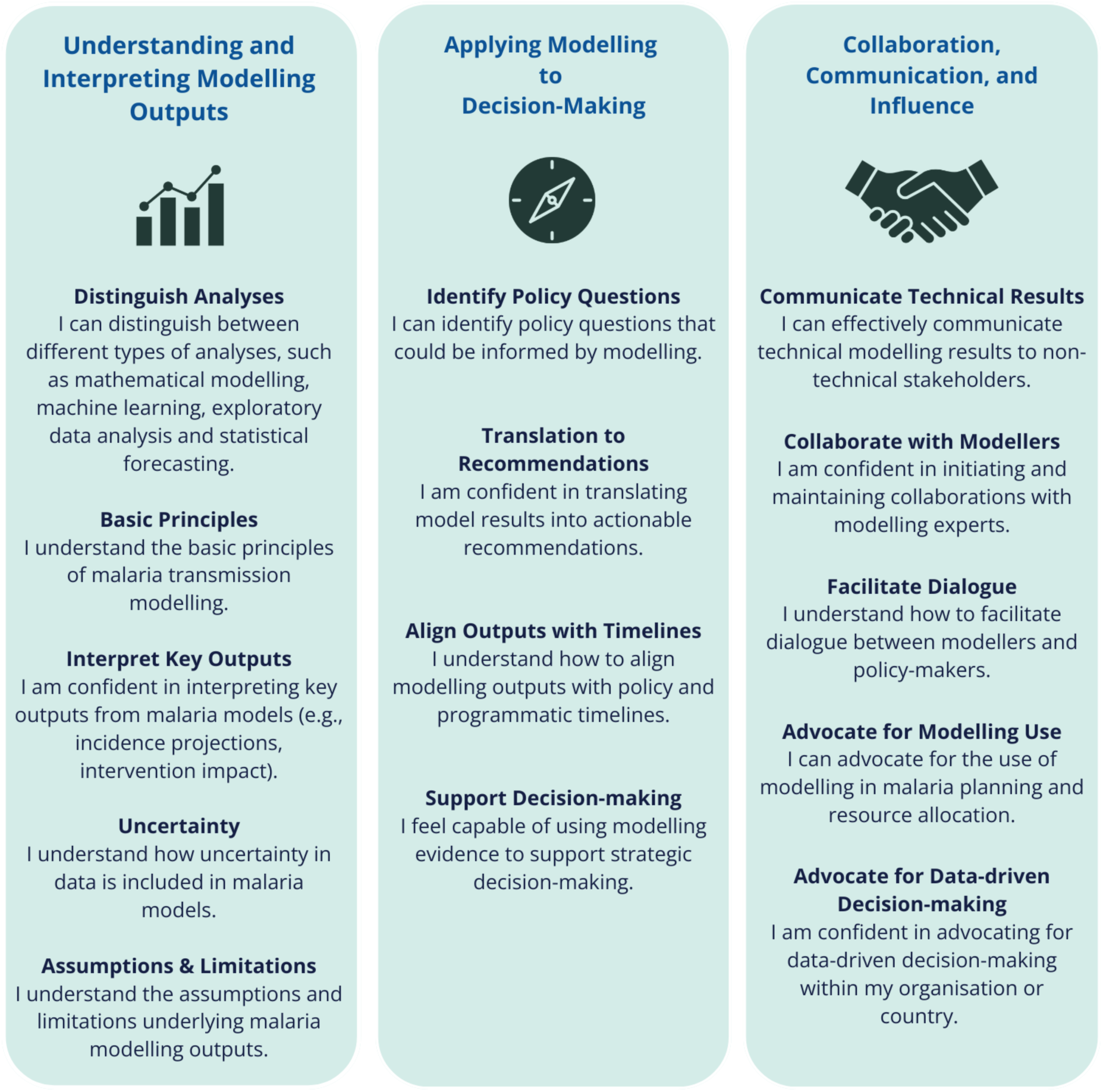
Competency Area and questions for the 14-item Likert scale

The 14 closed questions used a Likert 5-point scale—scored from 1 to 5, where 1 = Strongly Disagree, 2 = Disagree, 3 = Neutral, 4 = Agree, and 5 = Strongly Agree—and were the same throughout the three surveys. They were grouped into three themes: Understanding and interpreting modelling outputs; Applying modelling to decision-making; and Collaboration, communication, and influence. The open-ended questions varied across surveys.

## 3. Results

### 3.1 Curriculum

Table 1 presents the curriculum summary reflecting the balance between technical understanding with practical application to ensure fellows are equipped both to engage critically with models and use their results in real-world contexts. A detailed curriculum, complete with module guides, tools, teaching resources and facilitator guidance, is available online [6].

**Table 1.** Fellowship curriculum and learning outcomes mapped to competency domains.

|  | Topic | Domain* | Learning outcomes |
| --- | --- | --- | --- |
| 1 | Modelling:<br>What, Why and<br>How | 1, 2 | Understand what modelling is, and what purposes it serves in public health<br>Identify when modelling is an appropriate approach for informing decisions<br>Understand how different types of models are developed and applied |
| 2 | Systems<br>Thinking | 1 | Understand what systems thinking is, and how it applies to malaria programmes<br>Identify how interactions within complex systems influence disease outcomes<br>Explain how systems thinking can improve modelling and decision-making for malaria |
| 3 | Model Building<br>Process | 1, 2 | Identify the key steps in building a model from concept to application<br>Understand how assumptions, data, and structure combine within a model<br>Discuss the key principles of modelling for policy |
| 4 | Engaging with<br>Modelling | 2, 3 | Identify the factors that influence engagement with modelling in policy and programme settings<br>Understand the barriers that limit the uptake and use of modelling evidence |
|  |  |  | Articulate how funding and partnerships can support sustainable modelling efforts |
| 5 | Roles and Responsibilities | 2, 3 | Identify what roles different stakeholders play in the modelling process<br>Understand how collaboration between modellers and decision-makers can be structured effectively<br>Define responsibilities to support transparency and accountability in modelling |
| 6 | Question Formulation and Data Requirements | 1, 2 | Understand the kind of questions that modelling can address<br>Define clear, relevant, and actionable modelling questions<br>Identify what makes a modelling question useful for policy and decision-making<br>Evaluate common data sources needed to address different modelling questions and their suitability for modelling tasks |
| 7 | Designing Malaria Models | 1, 2 | Prioritise model features to address different malaria questions<br>Critically discuss the trade-offs exist between model complexity and usability<br>Understand how context influences model design choices |
| 8 | Critiquing Models | 1, 2 | Assess the strengths and limitations of a model<br>Review assumptions and outputs<br>Identify potential biases or misinterpretations |
| 9 | Data Visualisation and Interpretation | 1, 2 | Demonstrate effective ways to visualise modelling data and outputs<br>Understand how visualisation choices influence interpretation and decision-making<br>Identify common misinterpretations and communication challenges and how these can be avoided |
| 10 | Scenario Analysis | 1, 2 | Understand scenario analysis and how it is used in modelling<br>Design meaningful and realistic scenarios<br>Interpret and compare scenario results |
| 11 | Interpreting Model Outputs | 1, 2, 3 | Identify what model outputs represent, and how they should be interpreted<br>Explain how assumptions and uncertainty influence outputs<br>Demonstrate how outputs can be translated into actionable insights for decision-making |
| 12 | Uncertainty and Sensitivity Analysis | 1 | Explain what types of uncertainty exist in modelling<br>Understand how sensitivity analysis is used to assess the robustness of results<br>Demonstrate how uncertainty can be communicated effectively to non-technical audiences |
| 13 | Health Economics and Financing | 1, 2 | Explain how economic evidence can support policy and programme decisions<br>Identify what information is needed to assess affordability, sustainability, and value for money<br>Understand how modelling can inform funding allocations and investment strategies |
| 14 | Implementing Modelling Projects | 2, 3 | Demonstrate how a SWOT analysis can support the planning of a modelling project<br>Identify what internal and external factors influence successful implementation<br>Understand how risks and opportunities can be managed strategically |
| 15 | Communicating Model Findings | 2, 3 | Understand good practices for communicating model findings<br>Demonstrate how model communication can be tailored to different audiences<br>Identify what tools and formats best support clear and impactful messaging |

Notes: \*Competency domains: (1) Understanding and Interpreting Modelling Outputs; (2) Applying Modelling to Decision-Making; (3) Collaboration, Communication, and Influence

### 3.2 Evaluation of early outcomes and impact

#### 3.2.1 Understanding and Interpreting Modelling Outputs

The first theme, *Understanding and Interpreting Modelling Outputs,* focuses on the fellows’ technical literacy and ability to critically evaluate modelling outputs. This theme started with the lowest confidence (average score of 3.51) and had an average increase of +0.93 across the five questions. The competences with the biggest increase in confidence were *Q4: Uncertainty* (total change of +1.25) and *Q5: Assumptions & Limitations* (total change of 0.91), confirming a substantial increase in critical model literacy.

**Figure 4.**
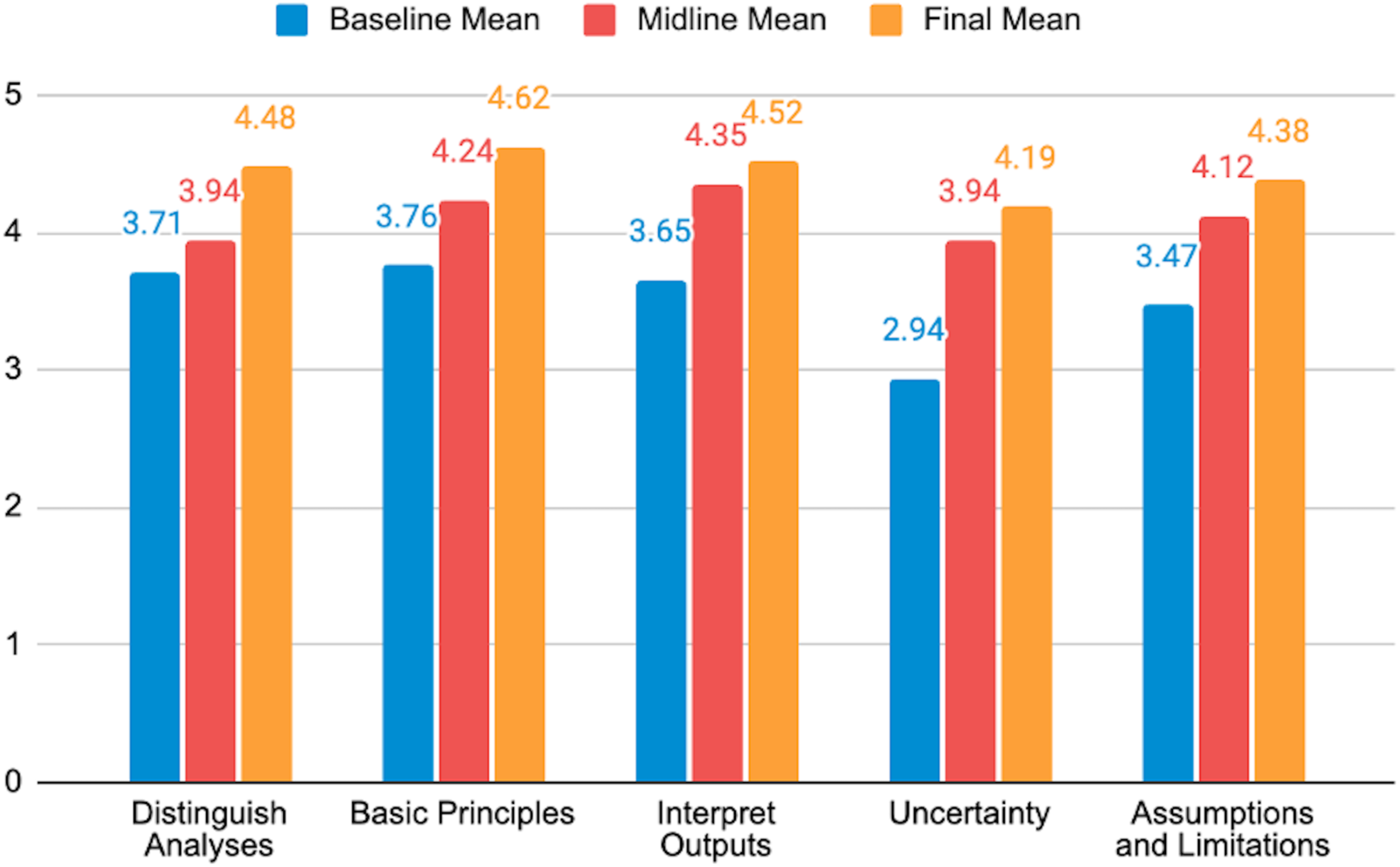
Survey results: Understanding and Interpreting Modelling Outputs

The final average score of 4.44 across these foundational competencies confirms fellows’ feelings of increased confidence in understanding not just the outputs, but also nuances and limitations of the models.

#### 3.2.2 Applying Modelling to Decision-Making

The *Applying Modelling to Decision-Making* theme assessed the fellows’ ability to translate technical knowledge into policy and strategic choices, representing the core objective of the Fellowship. It achieved the highest thematic growth with a total change of +0.97, moving from 3.46 at baseline to 4.43 as the final average score.

**Figure 5.**
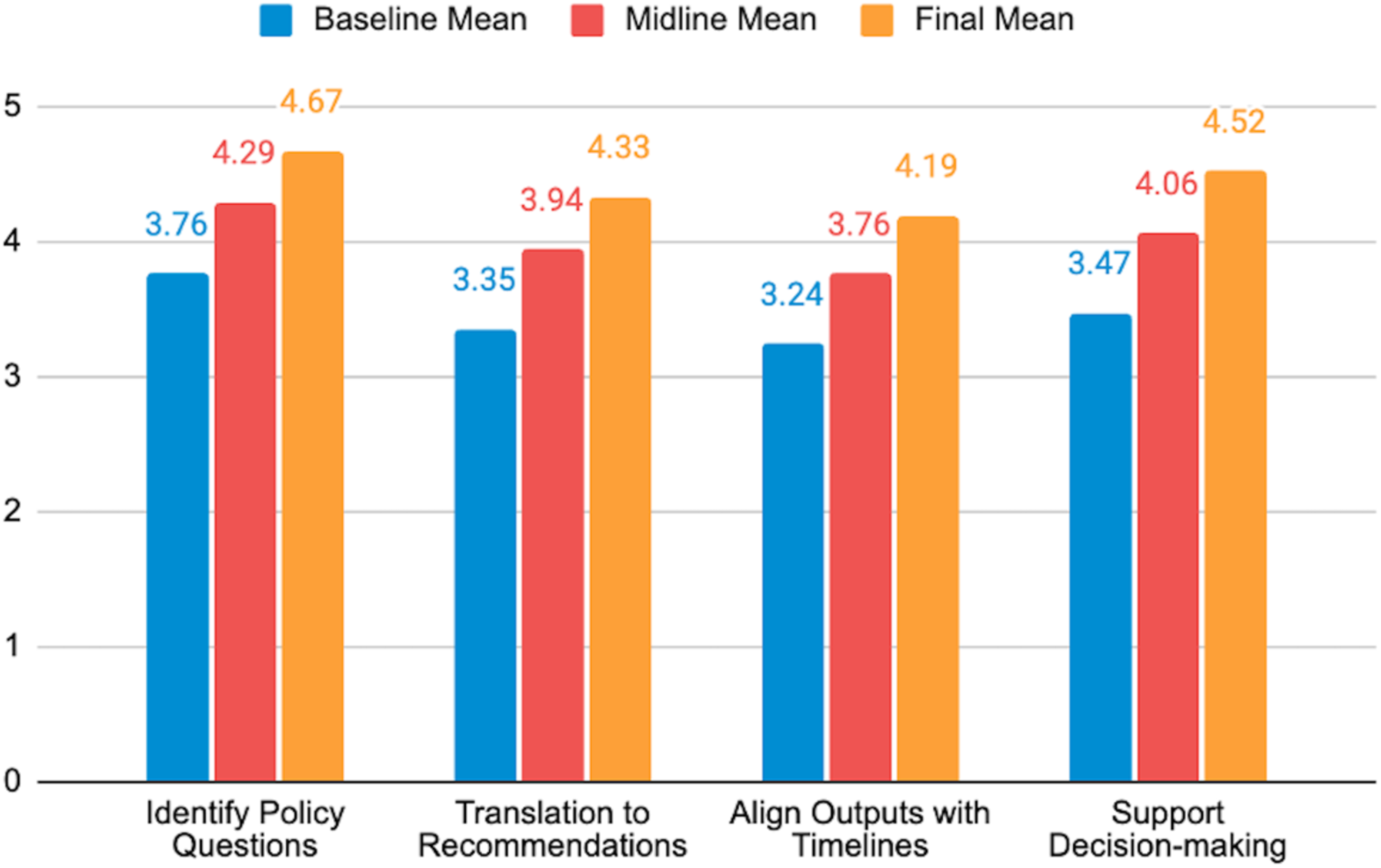
Survey results: Applying Modelling to Decision-Making

Two competencies in this theme showed an approximate full point increase: *Q9: Support Decision-making* (total change: +1.05) and *Q7: Translation to Recommendations* (total change: +0.98). This is an important finding for policymakers, demonstrating that fellows believe they are now better equipped to use modelling to drive strategic choices. The highest final mean was for *Q6: Identify Policy Questions* (average final score of 4.67) indicating that fellows are confident at defining the impactful questions for models to address, ensuring the evidence generated is relevant to policy needs.

#### 3.2.3 Collaboration, Communication, and Influence

The last theme, *Collaboration, Communication, and Influence*, captured the essential skills needed to bridge the policy-science divide and champion evidence-based decision-making. This theme concluded with the highest average final score of 4.49.

**Figure 6.**
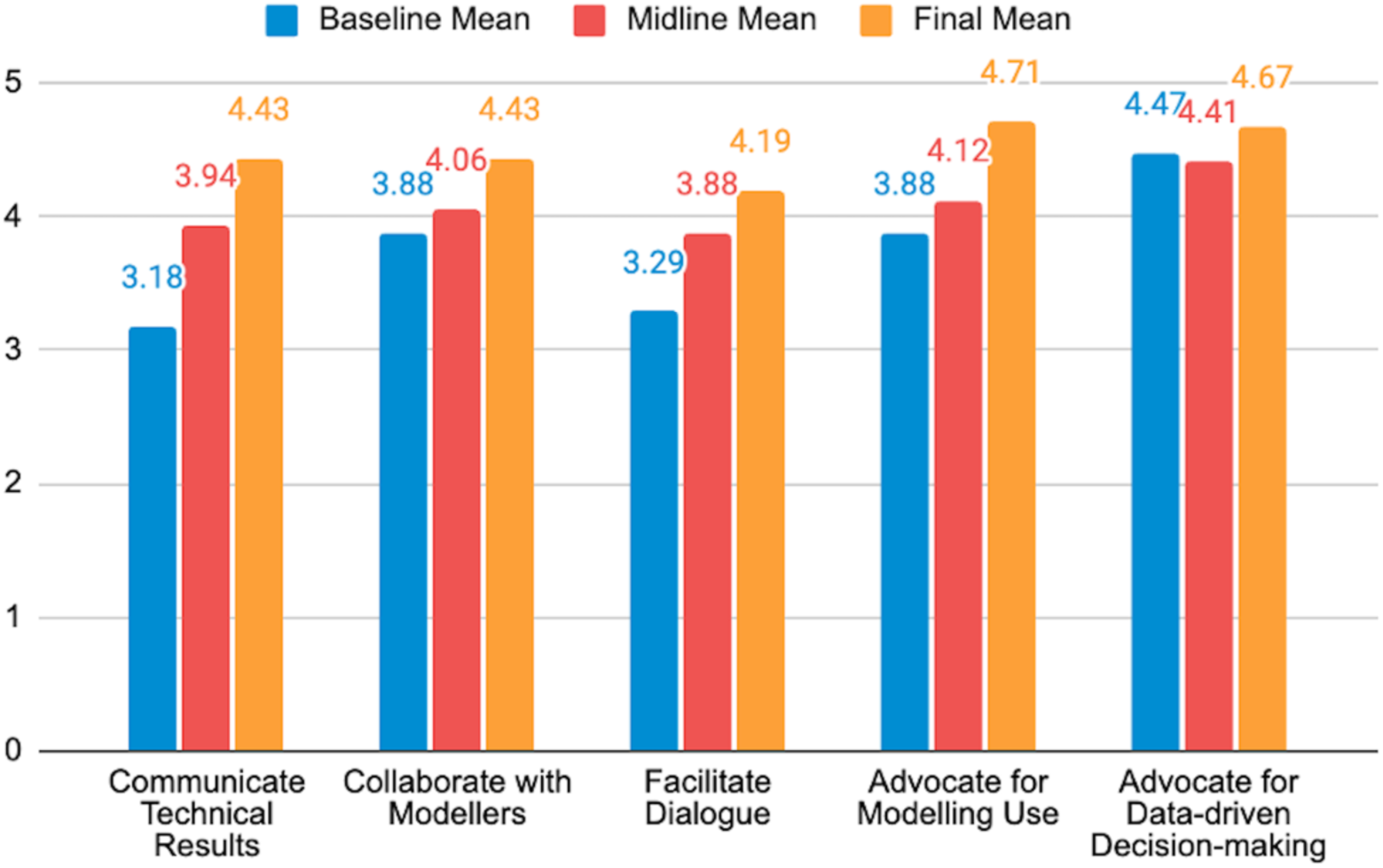
Survey results: Collaboration, Communication, and Influence

By the end of the fellowship, *Q10: Communicate Technical Results* (Total Change: +1.25) had one of the largest competency gains, demonstrating a substantial increase in the critical skill of translating complex technical information for non-technical stakeholders. There was also strong improvement in *Q12: Facilitate Dialogue* (Total Change: +0.90), indicating that fellows feel capable of managing constructive conversations between modellers and policy teams, fostering better collaboration and engagement. Fellow nominations prioritised those who were already working with data and making policy and implementation decisions. Thus, *Q14: Advocate for Data-Driven Decision-Making* had a high baseline (4.47) and was successfully maintained/slightly increased (4.67), reflecting a reinforced commitment to evidence use.

#### 3.2.4 Fellows’ Feedback

Based on the analysis of the open-ended questions in the final evaluation survey, the fellows’ perceptions of the RMMT Fellowship were positive, transformative, and action-oriented. The qualitative results align with the improvements seen in the quantitative evaluation, confirming the programme’s success in bridging the gap between technical modelling and policy application (Silal et al., 2026). Fellows consistently described the RMMT Fellowship as a highly impactful and often “transformative” experience that exceeded expectations, shifting their perception of modelling from a purely technical exercise to a practical and essential tool for policy and decision-making. A key theme was the strengthening of both technical and applied skills, including modelling processes, data analysis, scenario development, visualisation, and interpretation of results, alongside increased confidence in translating evidence into programmatic and policy-relevant recommendations. Fellows also highlighted a transformation in how they engage with modelling—moving from passive acceptance of results to critical evaluation of assumptions, outputs, and uncertainty.

The Fellowship’s emphasis on real-world application, hands-on learning, and policy relevance was widely valued, particularly in supporting strategic planning, resource allocation, and advocacy. In addition, fellows underscored the importance of collaboration and networking, noting that the programme fostered strong cross-country relationships and a growing community of practice. Overall, the Fellowship was seen as a catalyst for advancing data-driven decision-making, strengthening local capacity, and bridging the gap between modelling and policy in malaria programmes.

## 4. Discussion and Conclusion

### 4.1 Building translational modelling capacity

The Regional Malaria Modelling Translational (RMMT) Fellowship was developed to address a persistent challenge in infectious disease modelling: ensuring that increasingly sophisticated analytical methods are translated into evidence that is understood, trusted, and used to inform public health policy. While substantial investment has been made in advancing modelling methodologies, comparatively less attention has been directed towards strengthening the capacity of programme leaders and policymakers to engage critically with modelling evidence. Rather than training fellows to become modellers, the Fellowship sought to develop the skills required to commission, interpret, critique and communicate modelling analyses within decision-making processes. The improvements observed across all three competency domains, together with consistently positive qualitative feedback, suggest that this translational approach addresses an important and previously under-served capacity gap.

The Fellowship also demonstrated that translational capacity is built through sustained interaction between technical and policy communities. Throughout the programme, modelling was presented as a collaborative process requiring shared understanding between researchers, programme implementers, and policymakers. By emphasising question formulation, interpretation, uncertainty, and communication alongside modelling concepts, the curriculum positioned modelling as one component of an evidence-informed decision-making process rather than an isolated technical exercise.

### 4.2 Design features underpinning successful translational training

While the curriculum content was central to the Fellowship, several intentional programme design features contributed to fellow engagement and learning outcomes. A key design decision was recruitment through Ministries of Health rather than open individual application. Institutional nomination ensured organisational endorsement and encouraged countries to nominate fellows representing complementary leadership and technical roles. Bringing together programme managers, surveillance specialists, monitoring and evaluation officers, clinicians and data analysts created opportunities for peer learning across different perspectives while strengthening the likelihood that knowledge gained during the Fellowship could be translated into programme practice.

Institutional endorsement also helped fellows prioritise engagement throughout the six-month programme despite competing professional responsibilities.

Formal accreditation through the University of Cape Town further strengthened the Fellowship’s credibility. Positioning the programme as an accredited executive education course provided fellows with recognised professional development while reinforcing institutional confidence in the quality and rigour of the programme. Accreditation appeared to enhance fellow motivation and consistency of engagement, while signalling that translational modelling represents a recognised professional competency worthy of sustained investment.

The Fellowship’s long-format hybrid delivery model also emerged as an important educational strength. Combining immersive in-person teaching blocks with regular virtual sessions enabled fellows to revisit concepts over time, apply learning within their own institutional settings, and reflect on practical experiences between sessions. This allowed competencies to develop progressively. The improvements observed across the baseline, midline, and final evaluations are consistent with this iterative learning process and suggest that sustained engagement may be particularly valuable for executive education programmes where fellows are expected to apply learning within complex health systems.

Finally, the capstone projects anchored learning within nationally relevant policy priorities. Rather than reproducing technical modelling exercises, country teams developed proposals centred on real programme questions, considering epidemiological evidence, data requirements, health economics, implementation challenges, and stakeholder engagement. This approach reinforced two principles: (1) that modelling should begin with clearly defined policy questions and programme needs, ensuring that analytical methods remain driven by decision-making priorities rather than technical considerations alone, and (2) that decision-makers should be involved throughout the modelling exercise.

### 4.3 A replicable framework

Although developed within the context of malaria, the Fellowship was intentionally designed around competencies that extend beyond a single disease area. The educational framework focuses on identifying policy-relevant questions, understanding model assumptions and uncertainty, critically interpreting outputs, communicating evidence effectively, and facilitating collaboration between technical experts and decision-makers. These competencies underpin the effective use of modelling more broadly and therefore provide a strong foundation for adaptation to other public health programmes.

Transfer of the Fellowship to other disease areas would primarily require adaptation of epidemiological examples, modelling case studies, and policy frameworks while preserving the underlying educational principles and programme structure. The publication of the curriculum as an openly accessible educational resource is intended to facilitate such adaptation and encourage wider implementation.

The rapid evolution of artificial intelligence and increasingly accessible modelling platforms further reinforces the importance of this educational approach. As sophisticated analytical tools become available to a broader range of users, the principal challenge will increasingly shift from generating model outputs to evaluating whether those outputs are appropriate for specific policy questions. Future public health leaders will therefore require sufficient modelling literacy to critically assess assumptions, uncertainty, data quality, and limitations while engaging confidently with technical experts. Building translational capability alongside technical innovation will be essential to ensure that advances in modelling translate into meaningful improvements in public health decision-making.

### 4.4 Future directions for strengthening delivery and sustaining community

The inaugural Fellowship identified several opportunities to strengthen future iterations. Earlier identification of country modelling priorities could allow fellows to begin the programme with clearly defined policy questions, providing additional time for refinement of capstone projects and engagement with modelling teams. However, these questions will need to be assessed in advance so as to determine their suitability to modelling approaches. As countries continue to build modelling capacity, follow-on advanced modules could support fellows as projects progress from proposal development towards implementation and policy uptake.

The Fellowship has also established the foundations of a regional community of practice connecting policymakers, programme implementers, and modellers across participating countries. Sustaining these relationships beyond the duration of the Fellowship may be as important as the formal teaching programme itself, providing opportunities for continued collaboration, peer learning, and knowledge exchange. Building on the Fellowship, the preparations for the next phase are underway to support the inaugural cohort to implement their capstone projects through ongoing mentorship and technical collaboration.

In the longer term, embedding the curriculum within a modular postgraduate qualification or executive education pathway could provide a sustainable mechanism for continued professional development while strengthening institutional capacity across the region.

### 4.5 Limitations

Several limitations should be acknowledged. The evaluation reflects a single cohort of 21 fellows from seven countries and primarily measures self-reported confidence rather than objective assessments of competency or downstream policy impact. Although the capstone projects and qualitative feedback provide encouraging evidence of learning and application, longer-term evaluation is required to determine whether participation influences the routine commissioning, interpretation, and use of modelling within national malaria programmes and contributes to improved policy decisions.

The Fellowship was also intentionally resource-intensive, requiring substantial investment in faculty time, fellow support, travel, and residential teaching. While these elements contributed to fellow engagement and relationship building, they may limit scalability in some settings. Future implementation should therefore explore approaches that preserve the educational benefits of the Fellowship while increasing accessibility through regional partnerships, blended delivery models, institutional embedding, and train-the-trainer approaches.

### 4.6 Conclusion

The RMMT Fellowship demonstrates that strengthening the contribution of infectious disease modelling to public health requires investment not only in modelling methods but also in the people responsible for interpreting and applying modelling evidence. By combining institutional recruitment, academic accreditation, longitudinal hybrid learning, collaborative teaching, and policy-driven capstone projects, the Fellowship provides a practical and transferable framework for developing translational modelling capacity among programme leaders and decision-makers. Although developed for malaria, the educational principles underpinning the Fellowship are readily adaptable to other infectious disease programmes and public health contexts. As modelling methods, forecasting approaches, and artificial intelligence continue to evolve, building a workforce capable of critically engaging with increasingly sophisticated analytical tools will become essential for ensuring that modelling evidence is translated into effective policy and practice. The openly available curriculum provides a foundation to support this adaptation and encourages broader implementation, evaluation, and refinement of translational modelling education.

## Data Availability

All data produced in the present work are contained in the manuscript or available online at: https://uct-masha.github.io/RMMTFellowship/

## Acknowledgements

We acknowledge the leadership and collaboration of our implementing partners—Modelling and Simulation Hub, Africa (MASHA), University of Cape Town; SADC Malaria Elimination 8 Initiative (E8); Clinton Health Access Initiative (CHAI) South Africa Office—for their role in the design and delivery of the Fellowship. We extend our appreciation to the Ministries of Health and National Malaria Control Programmes in Angola, Eswatini, Ghana, Mozambique, Namibia, South Africa, and Zambia, for their engagement and support, and for enabling fellows to participate and apply their learning within national programmes. We recognise the contributions of the RMMT Fellows, whose commitment and engagement were central to the success of the programme, as well as the MMALA PhD cohort, whose participation strengthened linkages between modelling expertise and programme needs. We also thank the faculty, facilitators, and guest speakers, for their valuable contributions, and the programme coordination team, for their support in delivering the Fellowship.

## Author contributions

Conceptualization (SPS, RAH, SW); Methodology (SPS, RAH); Writing – original draft (SPS, RAH); Writing – review and editing (SPS, RAH, SW); Formal analysis (SPS, RAH, SW); Funding Acquisition (SPS). All authors approved the final manuscript.

## Data availability

All relevant data are publicly available.

